# Vestibulo-Ocular Reflex Function in Individuals with Car Sickness: A Cross-Sectional Study

**DOI:** 10.1101/2025.01.19.25320802

**Authors:** Oluwaseun I. Ambode, Eric G. Johnson, Pablo Mleziva, Summer San Lucas, Noha S. Daher

**Affiliations:** Department of Physical Therapy, Loma Linda University, CA, USA; Department of Allied Health Studies, Loma Linda University, CA, USA

**Keywords:** Chronic motion sensitivity (CMS), Bertec vision advantage (BVA), Vestibulo-ocular reflex (VOR), Postural stability, Physical activity

## Abstract

**Objective:** To determine whether Vestibulo-ocular reflex (VOR) integrity is different between young adults with and without car sickness, To assess the effect of gender on VOR integrity in young adults with and without car sickness and To compare postural stability across different levels of physical activity in young adults with and without car sickness.

**Design:** Cross-sectional study

**Setting:** General community

**Participants:** Forty-six healthy young adult men and women (ages 20-40 years) with and without car sickness

**Main Outcome measures:** Bertec Vision Advantage (BVA) was used to assess VOR integrity, computerized dynamic posturography with immersion virtual reality(CDP-IVR) was used to assess postural stability and the International Physical Activity Questionnaire (IPAQ-SF) was used to assess physical activity.

**Results:** Among females, there was no significant difference in BVA outcomes between the car sickness and no car sickness groups (p>0.05, Table 2), In males, there was no significant difference in median (Inter quartile range (IQR) outcomes by study group except for DVA lines lost. Male participants in the car sickness group had more lines lost compared to males in the no car sickness group.

**Conclusion:** Our findings showed that male participants with car sickness appeared to have a weaker VOR compared to females. Also, we found no significant interaction between physical activity and postural stability in young adults with and without car sickness. Future studies should consider assessing postural stability in the sitting position to better replicate the functional position of driving.

## Introduction

Car sickness is a type of motion sickness that is primarily attributed to a vehicle’s motion and results in nausea with dynamic driving styles characterized by higher accelerations [1]. Car sickness is a common type of motion sickness, that is potentially disabling to its sufferers [2]. One of the disorder’s distinctive characteristics is that passengers are more likely to be impacted because of the visual-vestibular imbalance that occurs when reading or using a computer screen in a moving vehicle. Furthermore, drivers are less susceptible to car sickness because they anticipate the motion of the vehicle, thus experiencing a reduced sensitivity to the expected versus discerned motions [1].

Empirical data shows that women are more likely than males to experience motion sickness, particularly when reading as a back seat passenger [2]. However, a previous body of work by Alharbi et al. [3] found no significant difference in motion sickness symptoms by gender [2]. Common symptoms of car sickness range from mild stomach aches and headaches to dizziness, nausea, and vomiting following exposure to certain car motions [4]. This discomfort can arise in other modes of transportation (such as ships, and buses) and is particularly relevant in the context of automated vehicles. According to the International Society of Automotive Engineers, self-driving vehicles present an opportunity for previous drivers to be relieved of tasks like steering the vehicle or supervising the surroundings [5]. During road transport, women are also more likely to report sickness than men, with a ratio of four to three [6].

The origin of motion sickness has been attributed to different theories. The most accepted of these theories is the sensory conflict theory which states that motion sensitivity arises when there is a conflict between the input systems; principally the visual and vestibular system [7]. Car sickness is a common type of motion sickness, potentially impairing its sufferers and is thought to be due to a sensory mismatch between the visual and vestibular system [2] although, the specific function of the VOR in this cascade has not been fully elucidated. Researchers reported that physical and sports activities can promote a rearrangement process that reduces susceptibility to motion sickness [8]. The authors of the current study speculated that an impaired VOR is implicated in car sickness.

Therefore, the objectives of this study were to 1) Determine whether VOR integrity is different between young adults with and without car sickness; 2) Assess the effect of gender on VOR integrity in young adults with and without car sickness, 3) Compare postural stability across different levels of physical activity in young adults with and without car sickness. The authors hypothesized that young adults with car sickness would demonstrate impaired VOR function compared to those without car sickness, VOR integrity would be influenced by gender in young adults with and without car sickness, and there would be a difference in mean postural stability by physical activity in young adults both with and without car sickness.

## Methods

### Participants

Fifty-nine participants between the ages of 20 and 40 with and without a self-reported history of car sickness were recruited using flyers, email, and word of mouth from the University and surrounding communities. The recruitment period for this study started on September 29^th^, 2023 and ended on February 2nd, 2024. Written informed consent was sought and obtain prior to data collection. The study was conducted in the Physical Therapy Department’s Neuroscience Research Laboratory. Participants were excluded if they reported a medical diagnosis of central nervous system disorder, impaired cervical spine range of motion, migraines, seizure disorder, vestibular dysfunction, or any musculoskeletal dysfunction that could limit their participation in the study. A total of 59 participants were assessed for eligibility. Thirteen were excluded due to technical difficulties and incomplete data, with the final sample size being 46 participants.

### Group Assignment

Participants were assigned to one of two groups: car sickness group or no car sickness group using the car sickness question on the Motion Sickness Susceptibility Questionnaire-Short Form (MSSQ-SF) [3,9]. The MSSQ-SF was used in a novel way. Participants who were sometimes or frequently sick while riding in cars were classified as adults with car sickness, and participants who never or rarely ill while riding in cars as adults with no car sickness. The validity and reliability of the MSSQ-SF have been demonstrated through its capacity to anticipate individual variances in CMS resulting from different motion types. The MSSQ-SF displayed strong internal consistency (Cronbach’s alpha = 0.87), good test-retest reliability (r= 0.9), moderate correlation between Section A (child) and Section B (adult) (r = 0.68), and predictive validity for motion susceptibility (r = 0.51) [10].

### Study Design

This was a cross-sectional study with participant stratification using the car sickness question on the MSSQ-SF into two groups: car sickness and no car sickness groups.

### Apparatus

The Bertec® Vision AdvantageTM (Bertec® Corporation, Columbus, Ohio, USA) was used to conduct Dynamic Visual Acuity (DVA) and Gaze Stability Testing (GST). These tests utilized a wireless inertial measurement assembly attached to the participants’ forehead with an elastic band. Data collected was then analyzed using the Bertec Balance Advantage^TM^ software [11,12].

The Logarithm of the Minimum Angle of Resolution (LogMAR) units is the unit of measurement for visual acuity. These units are utilized instead of traditional measures like the Snellen chart because they account for distance and are correlated with the traditional Snellen chart. A LogMAR value of zero indicates 20/20 vision, signifying normal vision. Lower LogMAR scores indicate better visual acuity, and a negative LogMAR rating denotes exceptional vision. Prior to conducting the DVAT and GST, baseline measurements for BLVA and VPT were obtained to customize the dynamic measures being assessed. The BLVA involves the presentation of the optotype “E” in various sizes to assess visual acuity with the head held still, whereas the VPT measures the time taken for the participant to identify the optotype “E” on the screen with a constant size. All passive head movements during the BVA were carried out by the primary investigator.

### Dynamic Visual Acuity Test

The DVAT assesses visual acuity by requiring continuous head movement of 20 degrees in each direction at a target velocity of approximately 100 degrees per second. The VPT results determine the visibility time of the optotype “E” on the screen. In the DVAT, the investigator guides participants in yaw plane head movement (left/right head rotation) at a target velocity of around 100 degrees per second with a range of 15 degrees above or below the target velocity. Throughout this movement, participants maintain their gaze on the screen and indicate the orientation of the optotype “E” (whether it is up, down, left, or right). As participants provide correct responses, the optotype becomes smaller and more challenging to identify. Conversely, incorrect responses lead to the optotype increasing in size, making it easier to identify the orientation. The lowest optotype level, where the participant accurately identifies the “E” orientation 60% of the time, marks the end of the examination and represents the final score. This process is carried out on both the right and left sides. The InVision System developed by Neurocom (Neurocom, Clackamas, Oregon, USA) showed moderate to good test-retest reliability in athletic populations with inter-rater reliability intra-class correlation coefficients (ICC) ranged from 0.323 to 0.937 for horizontal movements. The DVAT on the BVA is derived from data collected using the InVision System developed by Neurocom (Neurocom, Clackamas, Oregon, USA).

### Gaze Stability Test

The GST utilized the VPT value obtained at the beginning, the participant’s test direction (left or right), and the specific optotype set at a LogMAR level 0.2 higher than their BLVA. To conduct the GST, the researcher oscillated the participant’s head from left to right, reciprocally, within the yaw plane, with a range of 20 degrees in each direction. The software autonomously determined the starting direction for this evaluation, with a testing speed of approximately 80 degrees per second, not exceeding 150 degrees per second. The velocity was adjusted based on the participant’s ability to accurately identify the orientation of the optotype “E.” After being unable to accurately determine the orientation of the letter E, the highest rotational speed at which the direction of the “E” was correctly identified at a 60% standard was labeled as the GST score. This procedure was carried out on both the right and left sides.

### Computerized Dynamic Posturography with Immersion Virtual Reality

The Bertec Balance Advantage–Computerized Dynamic Posturography with Immersion Virtual Reality (CDP-IVR) from Bertec Advantage Corporation in Columbus, OH, USA was used to measure postural stability. Using the CDP-IVR, the participants’ postural sway and center of gravity displacement were assessed to generate an overall equilibrium score. The CDP-IVR analyzed and sampled signals from the participants’ attempts to maintain balance to determine postural sway and center of gravity displacement. This estimates equilibrium scores, which assessed how well participants’ sway remained within expected angular limits of stability during each testing condition. Those who exhibit minimal sway achieved equilibrium scores around 100, while participants with sway close to their stability limits scored close to zero [13].

### International Physical Activity Questionnaire (IPAQ-short form)

The IPAQ assesses physical activity in individuals aged 15 to 69 years old through self-reporting [14]. The assessment includes walking, moderate-intensity activities, and vigorous intensity activities, with a separate collection of frequencies (days per week) and durations (time per day) for each type of activity [15]. The IPAQ-SF is designed to capture physical activity based on a 7-day recall. The tool relies on self-assessment, categorizing participants as either inactive, minimally active, or engaging in health-enhancing physical activity (HEPA active) [14], or as having low, moderate, or high levels of physical activity [16]. Their is moderate correlation between self-reported; accelerometer-derived physical activity levels and the International Physical Activity Questionnaire-Short Form, with a high agreement level of 77.4%, and an intraclass correlation coefficient of 0.52–0.70 [17].

### Study Protocol

Following recruitment, study details were emailed before the laboratory meeting. The participant was briefed on the study at the laboratory, and biodata information, objective forms such as the International Physical Activity Questionnaire, and informed consent were collected. Participants were categorized into either the car sickness or no-car sickness group based on their response to the car sickness question on the MSSQ-SF scores. Both groups underwent assessment using the CDP-IVR under condition 1 (Dynamic vision, Stable platform) and condition 2 (Dynamic vision, Dynamic platform), as well as the BVA, which included VPT, BLVA, DVAT, and GST. The data was then synthesized and analyzed with inferences drawn.

### Data Analyses

Data was summarized using mean ± standard deviation (SD) for continuous variables, median (minimum, maximum) or median (interquartile range) for variables that are not approximately normal, and frequency (%) for categorical variables. The normality of the quantitative outcomes was examined using Shapiro test and boxplots. Participants’ characteristics were compared between the car sickness and no car sickness groups using independent t-test for quantitative variables, Mann-Whitney U test when the distribution of the variable was not approximately normal, and chi-square analysis for qualitative variables. Median and Interquartile Range (IQR) of BVA outcomes by study group were compared using Mann-Whitney U test. Mean±SD equilibrium score for Condition 1 and Condition 2 by physical activity was assessed using Analysis of Covariance to control for gender. The level of significance was set at significance was set at α=0.05.

## Results

The study sample included 46 participants (21 males and 25 females) with mean±SD age of 27.1±3.4 years and body mass index of 27.6±4.7 kg/m^2^. Half of the participants (50%) reported high physical activity, 15 reported moderate physical activity (32.6%), and 8 participants reported low physical activity (17.4%). Only 5 (10.9%) indicated having an ear infection. The median (minimum, maximum) of stress level was 6 (0, 9). There were no significant differences in participants’ characteristics between the car sickness and no car sickness groups (p>0.05) except for gender where a significantly higher proportion of car sickness participants were females compared to males (77.3% versus 22.7%, χ^2^=8.93, p=0.003; Table 1).

**Table 1.** Mean ± SD of Participants’ Characteristics by Study Group (N=46)

|  | **car Sickness (n<sub>1</sub>= 22)** | **No car Sickness (n<sub>2</sub>= 24)** | **p-value** |
| --- | --- | --- | --- |
| **Age, yr,** | 27.1±3.8 | 27.1±3.1 | 0.994 |
| **BMI (Kg/m<sup>2</sup>)** | 27.0 ± 5.3 | 28.1± 4.2 | 0.417 |
| **Gender, n(%)** |  |  |  |
| **Female** | 17 (77.3) | 8 (33.3) | **0.003** |
| **Male** | 5 (22.7) | 16 (66.7) |  |
| **Stress level** | 5.3 ± 2.7 | 5.2 ± 2.3 | 0.977 |
| **MSSQ-SF score\*** | 12.1 (2.0, 36.0) | 5.8 (0.0, 25.2) | **0.002** |
| **Had ear infections, n(%)** | 2 (9.1) | 3(12.5) | 0.543 |
| **Physical activity, n(%)** |  |  | 0.589 |
| **Low** | 5 (22.7) | 3 (12.5) |  |
| **Moderate** | 6 (27.3) | 9 (37.5) |  |
| **High** | 11 (50) | 12 (50) |  |

Abbreviations: SD, Standard Deviation; BMI, Body Mass Index; MSSQ-SF, Motion Sickness Susceptibility Questionnaire-Short Form; \*, Median (Minimum, Maximum)

Since there was a significant association between gender and car sickness, we compared median (IQR) of BVA outcomes by study group in females and males separately (Tables 2 & 3). Among females, there were no significant differences BLVA, VPT, DVAT, DVA lines lost and GST between the car sickness and no car sickness groups (p>0.05, Table 2. In males, there were no significant difference in median (IQR) outcomes by study group except for DVA lines lost to the right and left. Male participants in the car sickness group had more lines lost than in the no car sickness group (Right: 1.50 (1.0, 3.3) versus 0.80 (0.0, 2.5), Z=2.03, p=0.04; Left: 1.80 (0.8, 2.8) versus 0.80 (0.0, 2.5), Z= 2.16, p=0.032; Table 3)

**Table 2.** Median (IQR) Outcomes by Car Sickness for Females (N=25)

| | car sickness ($n_1= 17$) | No car Sickness ($n_2= 8$) | p-value |
| --- | --- | --- | --- |
| **BLVA (LogMAR)** | -0.13 (0.05) | -0.15 (0.11) | 0.440 |
| **VPT (ms)** | 30.00 (0.00) | 30.00 (0.00) | 0.669 |
| **DVAT-R (LogMAR)** | -0.05 (0.17) | -0.06 (1.04) | 0.511 |
| **DVAT-L (LogMAR)** | -0.05 (0.26) | -0.06 (0.06) | 0.262 |
| **DVA LL- R\*** | 1.00 (0.20, 6.30) | 1.25 (-0.50, 2.50) | 0.977 |
| **DVA LL- L\*** | 1.50 (0.20, 3.80) | 1.00 (0.30, 2.00) | 0.511 |
| **GST-R (°/sec)** | 120.00 (70.00) | 142.50 (30.00) | 0.262 |
| **GST-L (°/sec)** | 145.00 (70.00) | 142.50 (41.00) | 0.588 |

Abbreviations: IQR, Interquartile Range; BLVA, Baseline Visual Acuity; LogMAR, Logarithm of the Minimum Angle of Resolution; VPT, Visual Processing Time; ms, milliseconds; DVAT-R, Dynamic Visual Acuity Test- Right; DVAT-L, Dynamic Visual Acuity Test- Left; DVA LL- R\*; Dynamic Visual Acuity Lines Lost- Right, DVA LL- L\*; Dynamic Visual Acuity Lines Lost- Left, GST-R, Gaze Stability Test-Right; GST-L, Gaze Stability Test- Left; \*: Median (Minimum, Maximum)

**Table 3.** Median (IQR) Outcomes by Car Sickness for Males (N=21)

|  | **car sickness (n<sub>1</sub>= 5)** | **No car Sickness (n<sub>2</sub>= 16)** | **p-value** |
| --- | --- | --- | --- |
| **BLVA (LogMAR)** | -0.15 (0.08) | -0.13 (0.05) | 0.75 |
| **VPT (ms)** | 30 (0.00) | 30 (0.00) | 1.00 |
| **DVAT-R (LogMAR)** | 0.00 (0.17) | -0.07 (0.14) | 0.075 |
| **DVAT-L (LogMAR)** | -0.05 (0.14) | -0.06 (0.04) | 0.208 |
| **DVA LL -R\*** | 1.50 (1.0, 3.3) | 0.80 (0.0, 2.5) | **0.040** |
| **DVA LL- L\*** | 1.80 (0.8, 2.8) | 0.80 (0.0, 2.5) | **0.032** |
| **GST-R (°/sec)** | 125.00 (43) | 148.00 (34) | 0.075 |
| **GST-L (°/sec)** | 95.00 (55) | 147.50 (24) | 0.091 |

Abbreviations: IQR, Interquartile Range; BLVA, Baseline Visual Acuity; LogMAR, Logarithm of the Minimum Angle of Resolution; VPT, Visual Processing Time; ms, milliseconds; DVAT-R, Dynamic Visual Acuity Test- Right; DVAT-L, Dynamic Visual Acuity Test- Left; DVA LL- R\*; Dynamic Visual Acuity Lines Lost- Right, DVA LL- L\*; Dynamic Visual Acuity Lines Lost- Left, GST-R, Gaze Stability Test-Right; GST-L, Gaze Stability Test-Left; \*: Median (Minimum, Maximum)

The mean ± SD of equilibrium score for Conditions 1 and 2 by physical activity and study group after controlling for gender is shown in Table 4. There was no significant difference in mean equilibrium score for Conditions 1 and 2 between the car sickness and No car sickness groups (p=0.176 and p=0.486)). In addition, the changes in equilibrium score for Conditions 1 & 2 by study group did not differ by physical activity as determined by the non-significant interaction between car sickness and IPAQ (p=0.06 and p=0.329)

**Table 4.** Mean ± SD of Equilibrium Score for Conditions 1 and 2 by Physical Activity and Study Group, Controlled for Gender (N=46)

|  | Group |  |  | IPAQ |  |  |  |  |
| --- | --- | --- | --- | --- | --- | --- | --- | --- |
| | Car sickness (n<sub>1</sub>=22) | No car sickness (n<sub>2</sub>=24) | p-value ($\eta^2$) | Low (n<sub>1</sub>=8) | Moderate (n<sub>2</sub>=15) | High (n<sub>3</sub>=23) | p-value ($\eta^2$) | p-value ($\eta^2$) (Group x IPAQ) |
| **Condition 1** | 90.5 ± 2.9 | 91.5 ± 2.0 | 0.176 (0.05) | 90.6 ± 4.0 | 90.8 ± 2.0 | 91.3 ± 2.2 | 0.683 (0.02) | 0.06 (0.13) |
| **Condition 2** | 48.2 ± 23.5 | 47.6 ± 16.2 | 0.486 (0.01) | 52.0 ± 15.5 | 46.9 ± 22.3 | 47.1 ± 20.0 | 0.782 (0.01) | 0.329 (0.06) |

Abbreviations: SD, Standard Deviation; $\eta^2$ , Eta squared; IPAQ, International Physical Activity Questionnaire

## Discussion

Car sickness is a type of motion sickness that affects about 60% of the general population, primarily passengers. Half of these passengers demonstrate high susceptibility and symptoms [18]. Based on previous body of work by Gaikwad et al. [9], it had been established that gaze stability exercises improved postural stability through forced use of the VOR in individuals with CMS, and, as such, we theorized that VOR impairment would be a driving force behind CMS. We also theorized that individuals with car sickness would demonstrate a similar impairment in VOR as participants with CMS. The present investigation assessed the VOR integrity in individuals with car sickness in a cross-sectional study using the BVA which is effective and portable.

Firstly, there were no significant differences in participants’ characteristics between the car sickness and no car sickness groups (p>0.05), except for gender, where a significantly higher proportion of car sickness participants were females compared to males (77.3% versus 22.7%, χ^2^=8.93, p=0.003; Table 1), consistent with current evidence, (2,6) which shows a higher incidence of car sickness in females compared to males. Since there was a significant association between gender and car sickness, we compared median (IQR) of BVA outcomes by study group in females and males separately (Tables 2 & 3). Among females, there were no significant differences in BVA outcomes between the car sickness and no car sickness groups (p>0.05, Table 2).

In males, there were no significant differences in BVA outcomes by study group except for DVA lines lost to the right and left. Male participants in the car sickness group had more lines lost compared to the no car sickness group, indicating a weaker VOR. These findings were consistent with our hypothesis that participants with car sickness would demonstrate impaired VOR compared to participants without car sickness. In our study, male participants demonstrated impaired VOR based on one or more DVA lines lost compared to the no car sickness group. With our female participants with car sickness, there were no significant differences in BVA outcome variables when compared to the no car sickness group. This was contradictory to our hypothesis given the fact that we expected pronounced VOR impairment among the females with car sickness given a higher incidence in that group. To the best of our knowledge, our study is the first to make use of the BVA to assess VOR integrity in individuals with car sickness and as such there is a dearth of homogenous studies for comparison.

Secondly, we compared postural stability among different levels of physical activity in young adults with and without car sickness. There was no significant difference in mean equilibrium score for Conditions 1 and 2 between the car sickness and no car sickness groups (p=0.176 and p=0.486). This is consistent with current evidence [6], which showed no significant difference in equilibrium scores in participants with and without car sickness following postural stability assessment using a sway-referenced platform. In our study, one major factor that should be taken into consideration is the fact that postural responses were measured in standing whereas, symptoms exhibited during car sickness occur while in the sitting position. Therefore, assessment of postural responses in the sitting position in this population might yield different findings.

In addition, the changes in equilibrium score for Conditions 1 & 2 by study group did not differ by physical activity as determined by the non-significant interaction between car sickness and IPAQ. We theorize that this lack of significant interaction between study group and IPAQ score for Conditions 1 and 2 is due to the homogeneity of the participants in terms of their physical activity as majority (82.6%) of our participants were in the moderate/high activity levels on the IPAQ. We speculate that with a heterogenous physical activity group, we would see a significant interaction between study group and IPAQ scores given that regular physical and sporting activity has been shown to reduced motion sickness symptoms based on previous research [8].

### Study Limitations

The current study had limitations. First, a majority (82.6%) of our participants had physical activity levels in the moderate to high group and as such, our physical activity grouping was very homogenous. The authors feel this activity level homogeneity had resultant effects on our findings for VOR integrity and postural stability. Furthermore, results from this study is limited to the young adult’s age range of 20 to 40 years and cannot be generalized to the wider population of individuals who have car.

## Conclusion

Study findings showed that male participants with car sickness had weaker VOR compared to females. There was no significant interaction between physical activity and postural stability in young adults with and without car sickness. Future studies should consider assessing physical activity in a more heterogeneous physical activity group, as well as assessing postural stability in the sitting position to better replicate the functional position of driving.

## Data Availability

All relevant data are within the manuscript and its Supporting Information files.

## Abbreviations

CMS: Chronic motion sensitivity
BVA: Bertec vision advantage
CDP-IVR: Computerized dynamic posturography with immersion virtual reality
VOR: Vestibulo-ocular reflex
MSSQ-SF: Motion Sickness Susceptibility Questionnaire-Short Form
IPAQ-SF: International Physical Activity Questionnaire
BLVA: Baseline visual acuity
VPT: Visual processing time
DVAT: Dynamic visual acuity test
GST: Gaze stability test

## Acknowledgements

The authors are grateful to the participants for their participation and commitment to the study. The authors also thank the School of Allied Health Professions of Loma Linda University for their support of this research.

## References

1) Schmidt EA, Kuiper OX, Wolter S, Diels C, Bos JE. An international survey on the incidence and modulating factors of carsickness. Transp. Res. Part F. 2020 May 1;71:76–87.

2) Perrin P, Lion A, Bosser G, Gauchard G, Meistelman C. Motion sickness in rally car co-drivers. Aviat. Space Environ. Med. 2013 May 1;84(5):473–7.

3) Alharbi AA, Johnson EG, Albalwi AA, Daher NS, Cordett TK, Ambode OI, Alshehri FH. Effect of visual input on postural stability in young adults with chronic motion sensitivity: a controlled cross-sectional study J Vestib Res. 2017 Jan 1;27(4):225–31.

4) Dennison MS, Wisti AZ, D’Zmura M. Use of physiological signals to predict cybersickness. Displays. 2016 Sep 1;44:42–52.

5) Ramaioli C, Steinmetzer T, Brietzke A, Meyer P, Pham Xuan R, Schneider E, Gorges M. Assessment of vestibulo-ocular reflex and its adaptation during stop-and-go car rides in motion sickness susceptible passengers. Exp. Brain Res. 2023 Jun;241(6):1523–31.

6) Dida M, Guerraz M, Barraud PA, Cian C. Relationship between Car-Sickness Susceptibility and Postural Activity: Could the Re-Weighting Strategy between Signals from Different Body Sensors Be an Underlying Factor? J. Sens. 2024 Feb 6;24(4):1046.

7) Reason JT. Motion sickness adaptation: a neural mismatch model. JRSM. 1978 Nov;71(11):819–29

8) Caillet G, Bosser G, Gauchard GC, Chau N, Benamghar L, Perrin PP. Effect of sporting activity practice on susceptibility to motion sickness. Brain Res. Bull. 2006 Apr 14;69(3):288–93.

9) Gaikwad SB, Johnson EG, Nelson TC, Ambode OI, Albalwi AA, Alharbi AA, Daher NS. Effect of gaze stability exercises on chronic motion sensitivity: a randomized controlled trial. JNPT. 2018 Apr 1;42(2):72–9.

10) Golding JF. Predicting individual differences in motion sickness susceptibility by questionnaire. Pers Individ Dif. 2006 Jul 1;41(2):237–48.

11) Hoffer ME, Gottshall KR, Moore R, Balough BJ, Wester D. Characterizing and treating dizziness after mild head trauma. Otol Neurotol. 2004;25(2):135–8.

12) Quintana C, Heebner NR, Olson AD, Abt JP, Hoch MC. Sport-specific differences in dynamic visual acuity and gaze stabilization in division-I collegiate athletes. J Vestib Res. 2020;30(4):249–57.

13) Bertec Balance Advantage Quick Reference Manual PDF download [Internet]. [cited 2024 Oct 16]. Available from: https://www.manualslib.com/manual/2453213/Bertec-Balance-Advantage.html

14) International Physical Activity Questionnaire – Long Form. (2021). Retrieved 14 November 2021, from https://www.sralab.org/rehabilitation-measures/international-physical-activity-questionnaire-long-form

15) IPAQ Research Committee. Guidelines for data processing and analysis of the International Physical Activity Questionnaire (IPAQ)-short and long forms. http://www.ipaq.ki.se/scoring.pdf. 2005.

16) Cheng HL. A simple, easy-to-use spreadsheet for automatic scoring of the International Physical Activity Questionnaire (IPAQ) Short Form. RG. 2016.

17) Murphy JJ, Murphy MH, MacDonncha C, Murphy N, Nevill AM, Woods CB. Validity and reliability of three self-report instruments for assessing attainment of physical activity guidelines in university students. MPEES 2017 Jul 3;21(3):134–41.

18) Henry EH, Bougard C, Bourdin C, Bringoux L. Car sickness in real driving conditions: Effect of lateral acceleration and predictability reflected by physiological changes. Transp. Res. Part F. 2023 Aug 1;97:123–39

